## Supplementary Material (updated 30/4/25) for "How does policy modelling work in practice? A global analysis on the use of epidemiological modelling in health crises"

**Supplementary Material
Hadley et al., 2025**

**Contained in this Supplementary Material are the following:
S1: Study eligibility and ineligibility criteria
S2: Interview schedule
S3: Illustrative definitions for level of government interaction with modelling
S4: Results on recommendations for future practice (from interviewees)
S5: Deriving the classification framework for outbreak modelling and policy**

**S1: Study eligibility and ineligibility criteria**

The principal eligibility and ineligibility criteria used to recruit study participants was as follows:
Principal eligibility criteria: Participants will be epidemiological modellers who produced scientific advice to inform their country’s response to the Covid-19 pandemic AND the scientific advisors and policy/ decision-makers who received this scientific advice and contributed to managing their country’s outbreak response, between January 2020 and December 2022. In some countries, this requires membership of a government scientific advisory group; in other countries the key actors are less clear and primary research will be done to identify the full pool of suitable participants. Ineligibility criteria: Scientists, advisors, and policy/ decision makers who informed their country’s response to the Covid-19 pandemic but did not interact with epidemiological *modelling* (e.g., only with laboratory work, virology, medicine, field epidemiology, economics etc.).

**S2: Interview schedule**

The interview questions are appended at the end of this Supplementary Material. The first interview schedule was directed at modellers; the second at science advisors and policy/ decision makers (model users).

**S3: Illustrative definitions for the level of government interaction with modelling**

In the main text (Results 3.1), we present findings on the level of government interaction with modelling. The following illustrative definitions were used:

Level of government interaction

‘Low’ = NA. Not studied.

‘Medium’ = Medium level of government interaction, such as having a modeller on the relevant national science advisory committee or equivalent.

‘High’ = High level of government interaction, such as having a member of government embedded in the Modelling Committee and attending modelling meetings or equivalent.

‘Very High’ = Very high level of government interaction, where modelling and policy is extremely close-knit but modellers are still independent.

‘Embedded’ = The modelling committee is a full government entity and all major members are considered government employees. Note - this could be employees of a traditional government ministerial department or an arms-length agency (as an example, see the UK’s distinctions [[3]](https://www.zotero.org/google-docs/?mmimD1)).

Each country was cross-compared and countries were elevated or reduced based on in-depth comparative descriptions from interviewees and immersion in the wider literature to give a *relative* ranking. The above definitions are therefore illustrative.

**S4: Results on recommendations for future practice (from interviewees)**

Towards the end of each interview, interviewees were asked to discuss their key takeaways and recommendations for the future of outbreak modelling for policy. Full results are presented below and a summary is provided in the main text. Recommendations are split by those which were universal, and those which are context-specific: for countries with limited modelling capacity, for countries where modelling work was independent from government, and additional recommendations provided by policy/ decision makers and science advisors.

Universal key takeaways

- Tremendous progress on the use of modelling at scale in outbreak response, in many countries for the first time.
- DATA. “Be extremely close to the data.” “The crucial resource is data”. Almost all interviewees stressed data access and quality as essential enablers. For example, in one country interviewees acknowledged that the modellers had unique access for understanding the nuance of the data and extracting desired data for modelling questions: “We could go to the people working on the data and say ‘these are the things that we need’”. This enabled for example analysis on reinfections. Interviewees working in Hong Kong and mainland China also identified a need to work more closely with the ‘data generator’ and are now considering how to build this capacity for the future.
- Motivating data. Modelling also has a role in *motivating* data gathering. Key actors from Kenya and France for example identified a need to engage with their Ministry of Health to convey modelling needs (i.e. data collection).
- Geographic differences. Australian interviewees highlighted that externally reported data, e.g. from the Western Pacific or WHO, out of country can be misused. The work needs to be context dependent when decisions are made. Similar concerns were raised by key actors in New Zealand, where for example Māori communities faced ‘national’ rather than localised models and a lack of resources complicated datasets - from an equity perspective marginalised communities couldn’t benefit from the same level of surveillance for example. This was also echoed in Canada, given the lack of disaggregated data on First Nations, Inuit, and Metis peoples in different provinces and territories across the country.
- SYSTEMS. Funding and maintenance/ survival of Covid-19 systems. Almost all interviewees cited long-term funding for modelling teams and modelling-policy systems as an obvious constraint or enabler on future practices. Countries that developed modelling capacity rapidly during the pandemic had strong desire to maintain both the expertise and modelling-policy relationships. Interviewees reported that for example Canada, Kenya, and South Korea were working to maintain or evolve their Covid-19 modelling-policy systems post-pandemic, while at the time of interview, New Zealand and South Africa for example had been so far unsuccessful in sourcing long-term funding for modelling and formal modelling committees had disbanded. However there have been *informal* efforts in New Zealand (and elsewhere) to maintain collaboration between public health, policy/ decision makers, and academic modellers. One interviewee hypothesised that modelling capacity will be continued not through formal structures but through continued outbreak response.
- Legitimacy. One of the biggest reported strengths in Kenya was that the modelling consortium was assigned *within* the Ministry of Health. This gave modelling official placement and recognition throughout the pandemic. Other countries where modelling was external but had high government linkage also echoed this view.
- Structural change in the “inter-epidemic” period. Surveillance systems need to be improved not during emergency responses. Capacity and trust take time to build. It was agreed that now is the most crucial time for countries to strengthen their systems.^[1]^
- Skill development in periods of peacetime. Australian modellers noted that they are currently experimenting with and strengthening forecasting targets and visualisation techniques, without the pressure of an emergency response, as undoubtedly are many other teams around the world. Additionally, skills can be honed in all sorts of disease settings, not just high priority ones.
- TRUST AND COMMUNICATION. Strong communication between modellers and policy/ decision makers was routinely highlighted as paramount to successful relationships. For example, interviewees from one country cited the strength of their appointed convenor’s communication and brokerage on both the technical and knowledge translation side. Having an individual on board whose speciality is in communication was desired, even just to free up the time of other key actors.
- Trust takes time to build. It is important to have an *advocate* for modellers working with government in peacetime.
- Overbelieving modelling or its capacity or other misunderstandings. Overbelieved modelling can be more dangerous than no modelling - “once you predict something, people stop thinking about it”. Key actors in Japan and South Korea described misunderstandings between scenario modelling and forecasts, even among the public health sector, acknowledging that it is essential for the limitations of modelling to be carefully explained. This (limitations) was echoed by all modellers in the study. The misalignment between poor data and impact on outputs for example was not effectively communicated in one setting. Language barriers can also amplify confusion; interviewees noted that there is no different word for ‘projection’ and ‘forecast’ in either Korean or Japanese for example.
- INTERDISCIPLINARITY AND RELATIONSHIPS, taking a holistic view. Modelling primarily, or in many cases solely, focused on modelling for *health outcomes* (cases, deaths, hospitalisations etc.). Modellers acknowledged the very narrow metrics of measuring the impact of public health measures, based largely on “just keeping people out of hospital”. While this is absolutely critical work, the inability to model the socioeconomic impact of restrictions was a concern for many. In Hong Kong, modelling lacked economics and behavioural science considerations, and it was remarked that a sole focus on health-outcomes work may have contributed to higher vaccine hesitancy. Key actors from Kenya stressed that future outbreak and modelling response needs to have a *holistic* view: “Some people have gone hungry or had their lives messed up a bit because we couldn’t, we didn’t have the time to know or to be able to prioritise those things equally”. While social science and modelling collaborations were not in place for Covid-19 in Kenya, there have been changes to build on this moving forward - in the current Rift Valley Fever outbreak, modellers have been working with their colleagues in the School of Anthropology, consulting social scientists to inform their work.
- Economics. Following on from the above point, there was a mix of views on whether or not epidemiological modelling should explicitly model costs and if it is solely the job of senior government to see the different forms of science. A country where modelling did have an ear to Economics during Covid-19 was that of Australia - key actors acknowledged the importance of having developed relations with the Treasury to enable consideration of the economic impacts of both social restrictions and disease burden. The Australian Treasury’s interest in the use of modelling reportedly grew over time as disease severity fell, to understand for example social impacts of workplace absenteeism and related supply chain issues. Policy/ decision makers in Colombia also frequently requested modelling to integrate with economics. Modellers acknowledged that this is very difficult to do but a conversation that needs to be had - between epidemiological modellers and economic modellers. Modellers from South Korea were similarly vocal that modelling focused on *health*, not economics, mental health etc. “The modelling is correct in terms of minimising cases but overall we don’t know what is best for the country”. They acknowledged it was the job of the decision makers at least in South Korea in bringing together different streams of science but important for all parties to acknowledge the health-focus so that modelling is not overbelieved.

Specific recommendations from policy/ decision makers and science advisors

- DATA. Access to quality data - “models are only as strong as the data inputs”. Many of the policy/ decision makers and science advisors in the study recognised that data is critical for modelling. Poor intervention data was cited as a specific barrier in Peru; the Ministry of Health and the Peruvian CDC are working to improve their data sources.
- SYSTEMS. Capacity - the capacity for modelling in some countries is still lacking. We need to increase this, by providing secure jobs for modellers and training in countries with fewer experts. In Japan, physics and economics experts carried out some of the modelling activity but did not have training in the basics of infectious disease epidemiology; there is a need to have modelling-specific groups rather than relying on one team. An interviewee from Peru cited a need to integrate training in university settings - “they teach Mathematics and Computer Science but not these epidemiology things - we need to engage with universities to train more undergraduates in these methods”. Despite a regional interest in the use of modelling in Latin America and the Caribbean, there is a *lack of capacity to train* people to model, and this is most notable that only a few Latin American countries practice infectious disease modelling.
- Canadian key actors also acknowledged the unprecedented speed in which scientists but especially modellers had to operate - they propose to facilitate a stronger onboarding process, making it easier to hire modellers and those adjacently supporting modelling work.
- COMMUNICATION. Part of improving capacity is in improving communication. Countries should focus on facilitating communication between policy/ decision makers and modellers, with co-creation of the model questions. Improving risk communication is also vital to better response and adherence in outbreaks.
- Complex models were at times overpitched; there is a need for simple models in policy spheres.
- Brokering. It is critical to have an interpreter or translator (who may or may not be a modeller) within policymaking spheres to provide a space for modelling. Key actors in Canada and New Zealand both highlighted that it is useful to have someone who can provide a ‘key takeaway’ or ‘one-liner’ to synthesise information into practical and brief terms. Trusted academics that were comfortable sharing ballpark estimates with senior government in for example New Zealand proved very useful operationally. Other individuals also commented that modellers’ method of providing results should be more policy-friendly, in order to be accepted as useful by policy and decision makers.
- Certainty. Building on the above point, interviewees commented that it is critical to understand that policy and decision makers need to convey certainty in policy responses, which is inherently at odds with aspects of modelling. The environment of academia (and what is valued) obscures the importance of more emergency response and findings needed from studies - actors acknowledged the difference between academic modelling and what is important in policy.
- Retrospective verification of your own modelling is essential. “Many modellers console themselves by saying that my results can naturally be wrong and it was the best in that situation. But such a method can slow down improvement.” Politicians and stakeholders remarked that it is essential for modellers to retrospectively evaluate their own modelling and to share why things failed when they did - honesty between researchers and politicians is essential for progress.
- INTERDISCIPLINARITY AND RELATIONSHIPS. A lack of pre-existing relationships between modellers and policy/ decision makers meant it was difficult to establish rapport urgently in New Zealand. However the unique nature of New Zealand’s smaller community and head of Government being invested in science facilitated an informal communication style. Interviewees commented that perhaps a more formal bureaucratic structure would have been constricting - mobility comes with informality.
- Sharing scientific advice and government consensus documents regularly with the public. “Publishing routinely was absolutely the right thing to do”. It is democratic, enabled transparency, accountability, and “made our work better”.
- AND FINALLY, it is “important to recognise that mistakes will be made, that policymakers are human, that models are imperfect, but that ‘incremental gains’ can and should be made”.

Specific recommendations from countries with small or limited modelling capacity

- SYSTEMS. Capacity - it is important to have an existing body of people working in infectious disease epidemiology. In one country, interviewees described a situation where exactly the same people were drawn into the response for Covid-19 as for previous infectious disease emergencies, likened to a ‘wholesale transfer’ from HIV to Covid-19. It was crucial to have these existing relationships.
- There were numerous calls (both from modellers and policy/ decision makers) for more training of modellers and more standing capacity for modelling with secure jobs in peacetime. A modeller from Uganda remarked “it is frustrating in Uganda that I can’t even tell you what modellers we have!”. Activities to build a modelling community in Uganda have so far been unsuccessful; however the prominence of modelling for policy in Uganda is growing and senior Presidential Advisors have recently provided training for around 70 policy and decision makers on understanding modelling evidence.
- Review. While the addition of more modellers was not possible or not universally drawback-free in some countries, many individuals with small modelling capacity cited the desire for in-country peer review systems for emergency work. “What if we made mistakes that we weren’t aware of?”. The idea of peer-review was also discussed by some in larger modelling consortiums, for example via an external network of modellers to facilitate innovation and robustness of in-country modelling.
- A novel solution to capacity - Australia had insufficient capacity for multiple groups working on scenario modelling, but it was acknowledged that this strengthened the single model with skills and expertise coming together from multiple universities. Interestingly, the Australian national government also hired consulting companies at times; a unique solution to modelling capacity constraints. This was felt to be beneficial when applied to an area where existing modelling teams had less skills (for example in vaccine distribution). Teams working within and outside government were able to gainfully collaborate, sharing complementary knowledge and approaches to rapidly address urgent questions about national immunisation thresholds informed by a robust knowledge of delivery constraints.

Specific recommendations from countries where modelling sat ‘outside’ of government

- SYSTEMS. In the UK, the pre-pandemic plan was for most modelling to be carried out *within* government, run by those employees of the country’s public health agency who attended the “SPI-M-O” modelling committee, with academic members of the committee supporting and acting as a challenge function. However due to various reasons this was not the case, and modelling work was predominantly carried out by the academic groups themselves. Interviewees acknowledged a true ‘fence’ between those *inside* the UK government and those *outside*. It was commented that the planned system would have been strained anyway, with the model ‘inside’ and the modellers ‘outside’. There was discussion in two interviews on other European countries who pulled their academic modellers inside government, which at times had the unfortunate result of divorcing modellers from the rest of their community. Modellers in the UK enjoyed a much more coherent and supportive community with open efforts.
- Preserving relationships and protecting against evolution of political systems. Modellers relaying to a government committee rather than one or two government individuals has likely preserved future modelling-policy relationships under high government turnover in New Zealand.
- Funding and not having the money to resource a group. In the UK, interviewees highlighted one of the key mistakes as not being able to pay for the modelling expertise and people that the country needed. Modelling support was provided at goodwill under the overwhelming fear and shock that Covid-19 brought. It might not be possible for this goodwill to be repeated.
- COMMUNICATION. One-directional communication from policy to modelling early on was identified as a real barrier in some countries. Without a discursive relationship, modellers in these settings had no opportunity to understand what is actually being asked and what they were actually saying. It was not until two-way communication and invited co-creation of policy questions that modelling-policy proliferated in these countries.
- Further to the above, explicit information on what the government is trying to do was deemed very useful. No model is policy neutral - “all models have to have an assumption about policy in them, even if the policy is to do nothing”.
- Brokerage. Kenyan actors noted that there was unclear messaging and competing scientific advice before establishing a proper modelling committee and route for translation. There are now desires and aims to keep the modelling committee structure post-pandemic.
- INTERDISCIPLINARITY AND RELATIONSHIPS. Lastly, in countries where emergency modelling sat ‘outside’ of government, there is a need for more in-housing or at least *relationships* and formal *agreements* in place ahead of time, such as memorandums of understanding. For example, one interviewee noted that the role of evidence commissioning structures for disease control is currently being assessed nationally in Australia.

Throughout, we see four driving enablers to successful outbreak modelling and policy: **data**, **systems**, **communication**, and **interdisciplinarity/ relationships** (Figure R3).

**S5: Deriving the classification framework for outbreak modelling and policy**

In the main text, we use Ideal-Type Analysis to construct a classification framework (typology) for outbreak modelling and policy following the method of Stapley et al. [[4]](https://www.zotero.org/google-docs/?TLmAy9). Stapley et al. has been used broadly since its introduction in 2022, including in public health. Our typology aimed to answer the question: “What are the key contexts to consider when comparing shared experiences and recommendations on the integration of outbreak modelling in policy?”. Work was led by researchers LH and AT and began with familiarisation with the dataset and writing case reconstructions to summarise the relevant findings in each interview transcript.

The next step was to construct ideal types. Systematic consideration of each case construction and cross-comparisons allowed the researchers to detect trends across the dataset in terms of “participants’ whole accounts of their experiences or perspectives” [[4]](https://www.zotero.org/google-docs/?TLmAy9). Detailed analysis of interviewees’ beliefs and recommendations identified two key features linking shared experiences: the size/type of modelling infrastructure and level of government interaction with modelling. These are structural features but other ways to stratify the dataset were also explored and subsequently dropped if the categorisation was not a robust and rigorous representation of the whole dataset. Examples of typologies that were not taken forward included: sorting interviewees by degree of formality (those who mainly discussed formal collaborations in the interview vs informal collaborations); presence of or no identified relationship with the World Health Organization (incomplete data); temporal considerations (past vs present vs future outbreak modelling and policy); and diversity of collaboration with other disciplines vs modelling-centric collaborations (incomplete data). Some trial rankings were made for interviewees, but it was evident that it would be difficult to classify individuals by these metrics. However, categorising countries by ‘size/type of modelling infrastructure’ and ‘level of government interaction with modelling’ were robust to testing and experimentation across the whole dataset. These hence formed the basis of our classification framework.

Section 3.1 identified four types of national modelling infrastructure (Figure R1) and four levels of government interaction with modelling (Table R1), creating sixteen possible categories of shared experience (Supplementary Figure 1). Clustering similar experiences with re-review of individual transcripts and comparing transcripts on a case-by-case basis led the researchers to reduce this to five ‘ideal types’ or categories of shared experience (Supplementary Figure 2). This classification is made concrete with the addition of category descriptions, optimal cases (identifying the country best representing each category), and full list of examples from the study, together with testing and credibility check. The final classification is presented in Figure R2 of the main text and Supplementary Table 1 below.

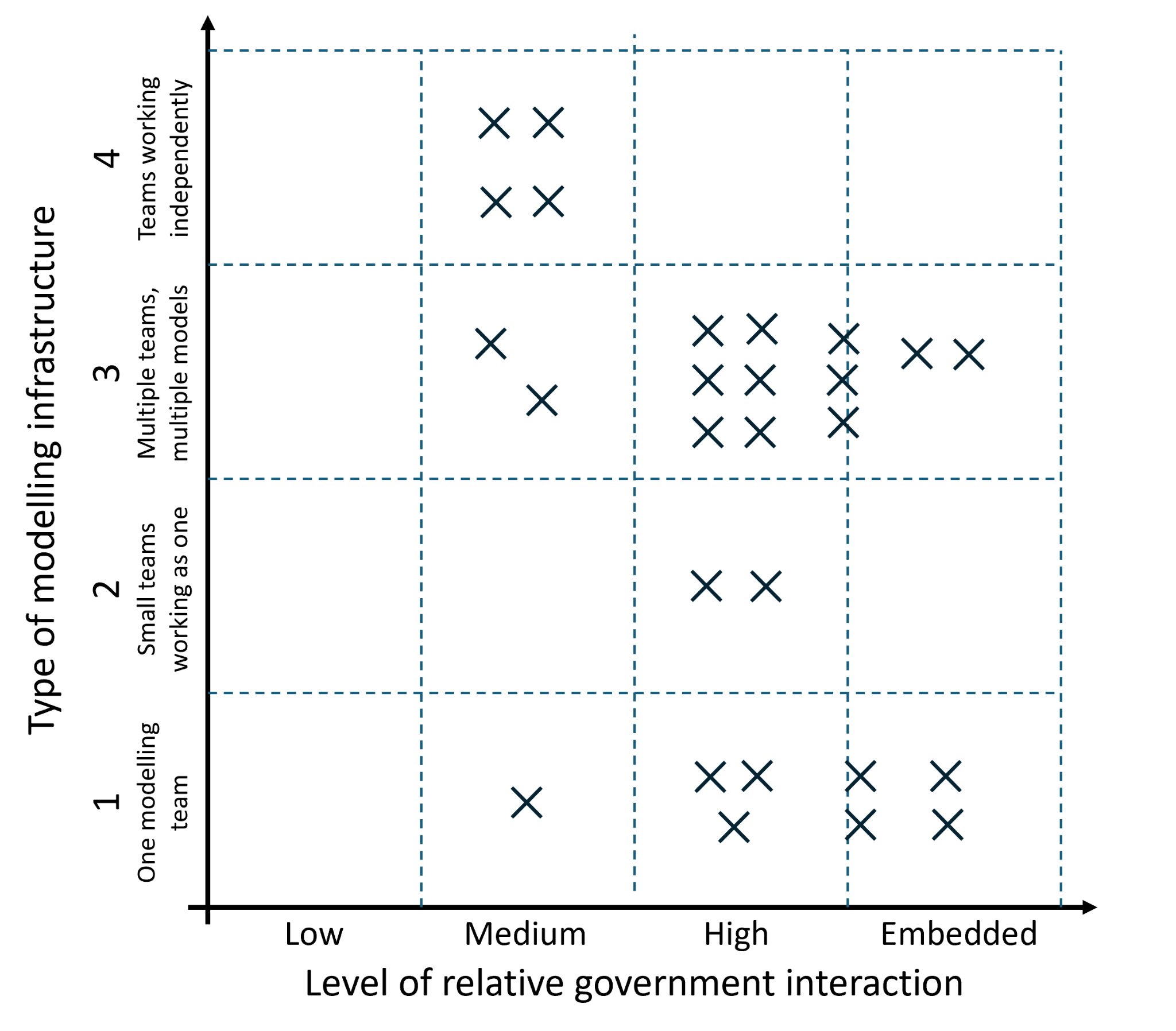

*Supplementary Figure 1: An early illustrative sketch plot for shared experiences, beliefs, and recommendations in outbreak modelling and policy. Points represent interviewees. Sketches such as these were used in preliminary Ideal-Type Analysis. Separate points were used for one interviewee in Australia who described scenario modelling and forecasting infrastructure separately. Case reconstructions for interviewees with ‘very high’ level of government involvement with modelling were re-analysed to explore if their experiences were similar to interviewees with government involvement rated ‘high’ or ‘embedded’ (or neither).*

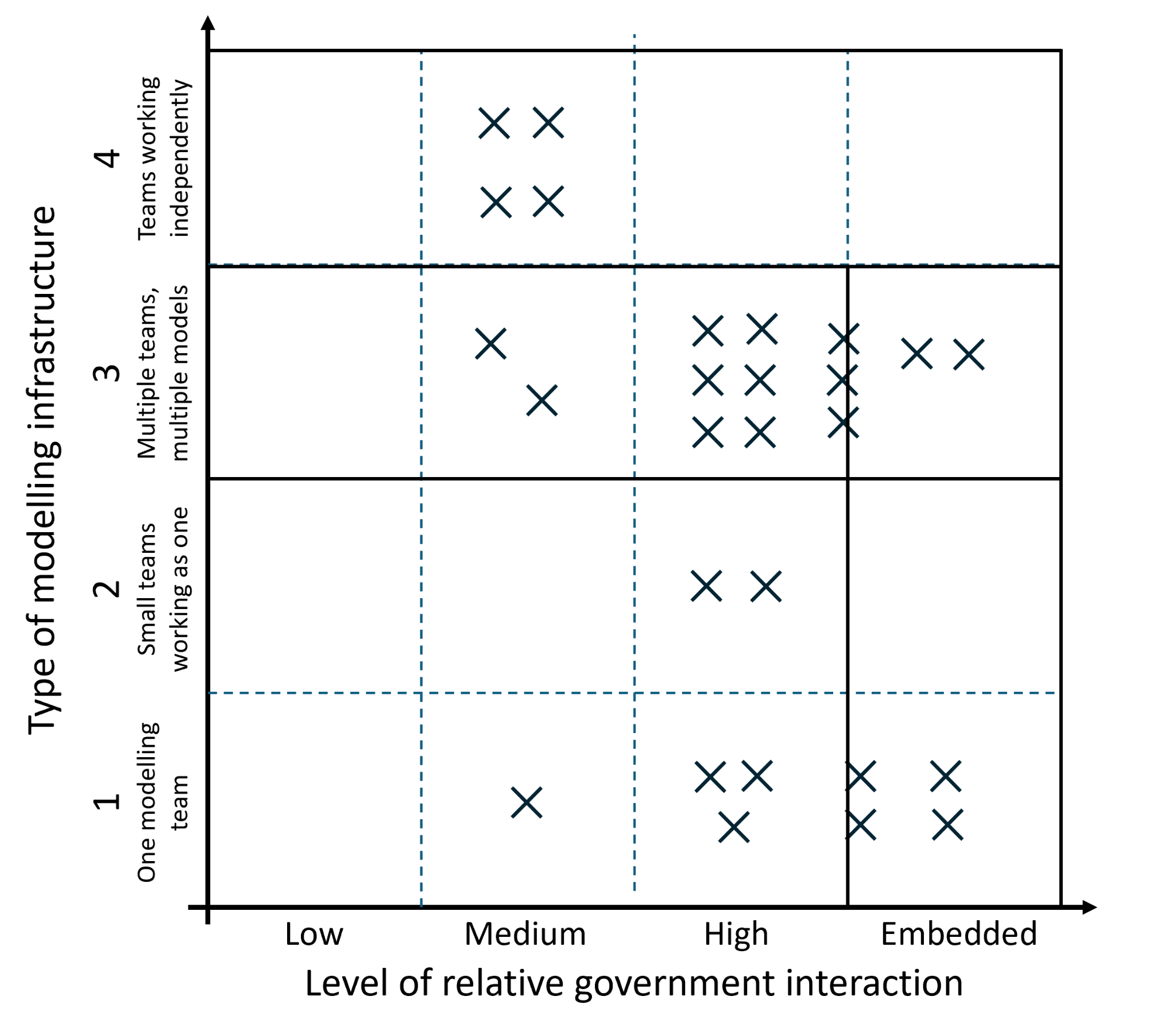

*Supplementary Figure 2: The illustrative sketch plot for shared experiences, beliefs, and recommendations in outbreak modelling and policy, from which categories of similar experience were derived. The five categories are marked out by solid black lines. As before, points represent interviewees. Sketches such as these were used in preliminary Ideal-Type Analysis.*

Category A (small modelling capacity, likely high government linkage):

Description -> “Low or focused modelling capacity (primarily one team and only one model) with little capacity to reproduce/validate, likely with high government linkage such as having government employees embedded in the modelling committee”.

Optimal case -> South Africa.

Examples in this cluster -> South Africa; Hong Kong; Australia (scenarios); France (in the early pandemic).

Category B (large modelling capacity, high to very high government linkage):

Description -> “Large modelling infrastructure (multiple teams, multiple models) with capacity to reproduce/verify, again with independence but likely with high government linkage such as (but not exclusively) having government employees embedded in the modelling committee”.

Optimal case -> Kenya.

Examples in this cluster -> Kenya; UK (in the later pandemic); Australia (forecasting); New Zealand; France (in the later pandemic).

Category C (teams working independently/ in isolation, combined by non-modellers in government):

Description -> “Modelling combined by government, where modelling teams/individuals are working independently, in isolation from each other. However knowledge transfer still occurs for example via a non-modelling SAC member”.

Optimal case -> South Korea.

Examples in this cluster -> South Korea; Uganda.

Category D (large modelling capacity with a primary in-government modelling team):

Description -> “Large modelling infrastructure (multiple teams, multiple models) with capacity to reproduce/verify, with a primary modelling team embedded in government”.

Optimal case -> Canada.

Examples in this cluster -> Canada; Colombia.

Category E (one in-government modelling team or similar):

Description -> “One government modelling team where modelling is done mostly or even exclusively within government. Modellers may be official government employees or could be researchers working very closely with government for the pandemic (pooling)”.

Optimal case -> Peru.

Examples in this cluster -> Peru; Japan.

*Supplementary Table 1: Final classification of shared experiences, beliefs, and recommendations in outbreak modelling and policy,* *as observed in our study.*

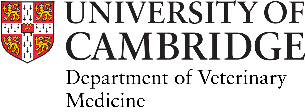

**Interview themes and questions**

**Study title**: “The use of infectious disease modelling in Covid-19 response: a multi-country study”.

We’ll start by sketching out the key actors together (via zoom whiteboard/ pen and paper), and then use the sketch and the below themes to guide our discussion. Opening questions are in blue; optional example follow-on questions in orange.

***Structures and pathways to policy***
*Could you sketch a rough diagram of ‘who’s who’ / who you feel was important in modelling and policy? (Illustrating some of the key actors and pathways in your country).*

- *Did you have a formal multi-model consortium/ ‘hub’, OR other formal pathway, OR no formal structures?*
- *How did the collaboration with policy begin? Who initiated?*
- *Any evolution of the structure through the pandemic?*
- *Training and certification: ‘Who’s in’? Whose advice is ‘trusted’?*
- *What worked? Recommendations? Practical and theoretical (tried/untried) suggestions welcome.*

***Collaboration and knowledge transfer***
*We’re interested in international knowledge transfer among modellers. Do you feel you had access to sufficient in-house expertise for modelling? Did you collaborate with other teams?*

- *Sharing of knowledge/ experiences/ contacts/ data/ models across countries?*
- *Strong national or international network of modellers? Would this be helpful?*
- *Thinking across disciplines. Did you have discussions with other scientists informing the response or were efforts quite disjoint? Economics/ behavioural science/ physicians etc.*
- *What worked? Recommendations?*

***Communication and visualisation***
*First thinking about communication: what level of discourse occurred between modellers and decision-makers throughout the pandemic?*

- *Translation. Who took up the role of translation? (Modellers/ Decision makers/ Intermediary/ Certain individuals). Do you feel this was done effectively?*
- *Visualisation. Can you give an example of a plot that you felt worked really well? And an example of one that was perhaps misunderstood or not quite so effective at demonstrating model findings? Wider question: what types of graphs and plots and reports were most helpful to decision-makers?*
- *Selective publishing. How do you decide what to include or exclude in reports/presentations to policy? Level of detail, how accurate?*

***Evaluation and reflection***
*What was your biggest mistake? Can you give an example of where you feel modelling got it wrong?*

- *Has there been interest in reflecting on how modelling can be made more useful to public health decision-makers? Perhaps through improved pathways, improved translation, and so on.*
- ***Lessons learned / your key takeaways from working on Covid?***
- *Future of modelling. What do you see the future of modelling for outbreaks to look like? How might we get there?*

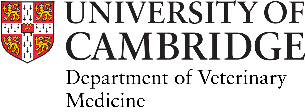

**Interview themes and questions**

**Study title**: “The use of infectious disease modelling in Covid-19 response: a multi-country study”.

The themes and questions below will be used as prompts to guide our interview discussion. Opening questions are in blue, example follow-on questions in orange. There will also be ample opportunity to discuss any other information that you feel is relevant, helping us to understand the interaction of modelling and policy during Covid-19 in your country of practice.

***Structures and pathways to policy***
*How did the collaboration with modelling begin? Is this the first time infectious disease modelling has been used to advise the government in your country?*

- *Do you feel that the setup was effective? Was it easy for modelling results to reach senior government? And could policymakers easily suggest questions of interest for modelling?*

***Communication and visualisation***
*What level of communication occurred between yourself and senior government, and yourself and modellers throughout the pandemic? For example, did you have the opportunity to discuss/report model findings with senior government on a regular basis and to feedback to the modelling teams?*

- *Translation. Who took up the role of translation (converting complex epidemiological findings into meaningful language for decision makers)?*
- *Visualisation. What aspects of graphs and plots were most helpful to decision-makers and advisors?*
- *Can you give an example of a plot that you felt worked well? And an example of one that was perhaps misunderstood by policymakers or not quite so effective at demonstrating model findings?*
- *Selective publishing. How do you decide what to include or exclude in reports/presentations to government? Level of detail? How accurate do you choose to be?*

***Collaboration and knowledge transfer***
*Do you feel you had access to sufficient country expertise for infectious disease modelling?*

- *Cross-disciplinarity. Were modellers encouraged to work closely with other scientists informing the response or were the different streams of evidence kept deliberately distinct? Economics/ behavioural science/ medicine etc.*

***Evaluation and reflection***
*What are your key takeaways or lessons learned from working in Covid-19 and supporting modelling? What was your biggest mistake (in relation to modelling)?*

- *Can you give an example of where you feel modelling got it wrong?*
- *Has there been interest in reflecting on how modelling can be made more useful to public health decision-makers? Perhaps through improved pathways, improved translation, and so on.*
- *Future of modelling. Do you think modelling will be used in future outbreaks in your country? What advice would you give to future actors?*

1. The authors note that not every country is in relative ‘peacetime’ at the time of writing this manuscript - one African interviewee stressed that “infectious diseases are a real and present danger in African settings in a way that they are not in most of the Global North”.
